# SARS-CoV-2 N-antigenemia: A new COVID-19 marker and a potential alternative to nucleic acid amplification techniques

**DOI:** 10.1101/2020.09.14.20191759

**Authors:** Quentin Le Hingrat, Benoit Visseaux, Cédric Laouenan, Sarah Tubiana, Lila Bouadma, Yazdan Yazdanpanah, Xavier Duval, Houria Ichou, Florence Damond, Mélanie Bertine, Nabil Benmalek, French COVID cohort management committee, CoV-CONTACT study group, Christophe Choquet, Jean-François Timsit, Jade Ghosn, Charlotte Charpentier, Diane Descamps, Nadhira Houhou-Fidouh

## Abstract

**Background:** Molecular assays on nasopharyngeal swabs remain the cornerstone of COVID-19 diagnostic. Despite massive worldwide efforts, the high technicalities of nasopharyngeal sampling and molecular assays, as well as scarce resources of reagents, limit our testing capabilities. Several strategies failed, to date, to fully alleviate this testing process (e.g. saliva sampling or antigen testing on nasopharyngeal samples). We assessed the performances of a new ELISA microplate assay quantifying SARS-CoV-2 nucleocapsid antigen (N-antigen) in serum or plasma.

**Methods:** The specificity of the assay, determined on 63 non-COVID patients, was 98.4% (95% confidence interval [CI], 85.3 to 100). Performances were determined on 227 serum samples from 165 patients with RT-PCR confirmed SARS-CoV-2 infection included in the French COVID and CoV-CONTACT cohorts.

**Findings:** Sensitivity was 132/142, 93.0% (95% CI, 84.7 to 100), within the first two weeks after symptoms onset. A subset of 73 COVID-19 patients had a serum collected within 24 hours following or preceding a positive nasopharyngeal swab. Among patients with high nasopharyngeal viral loads, Ct value below 30 and 33, only 1/50 and 4/67 tested negative for N-antigenemia, respectively. Among patients with a negative nasopharyngeal RT-PCR, 8/12 presented positive N-antigenemia. The lower respiratory tract was explored for 6/8 patients, showing positive PCR in 5 cases.

**Interpretation:** This is the first demonstration of the N-antigen antigenemia during COVID-19. Its detection presented a robust sensitivity, especially within the first 14 days after symptoms onset and high nasopharyngeal viral loads. These findings have to be confirmed with higher representation of outpatients. This approach could provide a valuable new option for COVID-19 diagnosis, only requiring a blood draw and easily scalable in all clinical laboratories.

## INTRODUCTION

Molecular assays on nasopharyngeal swabs remain the cornerstone of COVID-19 diagnostic. Despite massive worldwide efforts, the high technicalities of nasopharyngeal sampling and molecular assays, as well as scarce resources of reagents, limit our testing capabilities. Several strategies failed, to date, to fully alleviate this testing process *(e.g*. saliva sampling^1,2^ or antigen testing on nasopharyngeal samples^3^). To date, antigen detection has been only designed as point of care testing for nasopharyngeal samples with poor sensitivity^4^. No evaluation of N-antigen detection in other biological matrices are available.

In this work, we assessed the performances of N-antigen detection in sera, using a new and being marketed ELISA microplate assay, the COVID-19 Quantigene® (AAZ France), quantifying SARS-CoV-2 nucleocapsid antigen (N-antigen) in serum or plasma.

## METHODS

### Patients and ethics

Negative samples are composed of 50 pre-pandemic samples (collected between December 2, 2019 and January 13, 2020) and 13 pandemic samples from SARS-CoV-2 non-infected patients positive for other microbial antigens *(i.e*. NS1 antigen, HBs antigen, HIV-1 p24 antigen, HKU1 coronavirus, malaria antigens). Positive samples were collected between January 25, 2020 and September 2, 2020 from study participants included in the French COVID (clinicaltrials.gov NCT04262921) and CoV-CONTACT cohorts (clinicaltrials.gov NCT04259892). They have provided written informed consent for the use of their samples for research. Ethics approval was given by the French Ethics Committee CPP-Ile-de-France 6 (ID RCB: 2020-A00256-33 and ID RCB: 2020-A00280-39) and the French National Data Protection Commission (approval #920102).

### N-antigen levels assessment

Prior to analysis, sera samples were stored at −80°C. All samples were collected from March to June 2020. N antigenemia levels were determined with a being marketed CE-IVD ELISA microplate assay, COVID-Quantigene® (AAZ, Boulogne-Billancourt, France), according to manufacturer recommendations. Briefly, in each well of 96-wells microplates previously coated with anti-SARS-CoV-2 N-antibodies, 50 μl of a solution containing a biotinylated anti-SARS-CoV-2 N antibodies and 50 μl of sera were added. After incubation at 37°C for 60 minutes, plates were washed 5 times with a phosphate buffer solution. Then, 100 μl of a solution containing HRP-conjugated streptavidin were added, followed by incubation for 30 minutes at 37°C. Plates were washed 5 times with the phosphate buffer solution, then 50 μl of a solution containing the substrate (3,3’,5,5’-tetramethylbenzidine (TMB)) and 50 μl of a second solution containing urea were added. After 15 minutes at 37°C, the colorimetric reaction was stopped by adding 50 μl of H_2_SO_4_. Absorbance values were measured at 450nm, with a reference set at 630nm. In each plate, standards made of recombinant N antigens were tested, to quantify the N antigenemia levels for each patient’s sample. As the purpose of this study was to assess the sensitivity of this new assay, samples with titers above 180 pg/mL were not diluted for precise quantification.

### RT-PCR assays

For all patients included in this study, diagnosis of SARS-CoV-2 infection was performed in the virology department of Bichat-Claude Bernard University Hospital by RT-PCR on naso-pharyngeal swabs, as recommended.

Different techniques were performed throughout the study period for nasopharyngeal samples, due to frequent shortages issues and requirements for fast turnaround time: RealStar® SARS-CoV-2 (Altona, Germany), Cobas® SARS-CoV-2 (Roche, Switzerland), Simplexa® COVID-19 Direct (DiaSorin, Italy), BioFire® SARS-CoV-2 (BioMerieux, France), QIAstat-Dx® Respiratory SARS-CoV-2 (Qiagen, Germany) and NeumoDX® (QIAgen, Germany) using the IP2 Institute Pasteur and the WHO E gene primers^5^. As E gene was amplified by all techniques, except Simplexa® COVID-19 Direct, its cycle threshold (Ct) value was recorded.

For a subset of 146 sera samples, corresponding to 89 patients included in the French COVID-19 cohort, concomitant plasma samples were available, presence of viral RNA in plasma was determined. Briefly, viral nucleic acids were extracted from 200 μL of plasma with the MagNA Pure LC Total Nucleic Acid Isolation Kit - Large Volume (Roche Diagnostics) and eluted in 50 μL;. RT-PCR was performed on 10 μl of eluate using the RealStar® SARS-CoV-2 assay (Altona, Germany), according to the manufacturer recommendations. Samples with RT-PCR cycle threshold values above 40 were considered negative.

### Detection of anti-SARS-CoV-2 nucleocapsid IgG

For a subset of 85 sera, corresponding to 80 patients (ICU patients: n = 21, ward patients: n = 36 and outpatients: n = 36), we performed a chemiluminescent microparticle immunoassay detecting anti-N immunoglobulins G (Architect SARS-CoV-2 IG Assay, Abbott). Results were reported as a signal to cut-off (S/Co) value. The positivity threshold was set to 1.4, as recommended by the manufacturer.

### Data availability

A file compiling all data used in this article is available on Mendeley Data public repository (https://data.mendeley.eom/datasets/fiz6zbkxvm/1).

## RESULTS

We assessed the performances of a new ELISA microplate assay, the COVID-19 Quantigene® (AAZ France), quantifying SARS-CoV-2 nucleocapsid antigen (N-antigen) in serum or plasma. The specificity of this assay, determined on 63 non-COVID patients, was 98.4% (95% confidence interval [CI], 85.3 to 100). Performances for N-antigen detection was determined on 227 serum samples from 167 patients with RT-PCR confirmed SARS-CoV-2 infection included in the French COVID and CoV-CONTACT cohorts. Sensitivity was 132/142, 93.0% (95% CI, 84.7 to 100), within the first two weeks after symptoms onset (Figures 1A and 1B). Above 14 days from symptoms onset, antigenemia frequently declined, in link with anti-N IgG detection (figure 1A and Supplementary Appendix 1A), and was undetectable in 11/13 (84.6%) outpatients, 8/19 (42.1%) ward patients and 16/49 (32.7%) ICU patients. Patients with positive RNAaemia (viremic patients) exhibited higher N-antigen sera levels (Figure 1-C).

**Figure 1:**
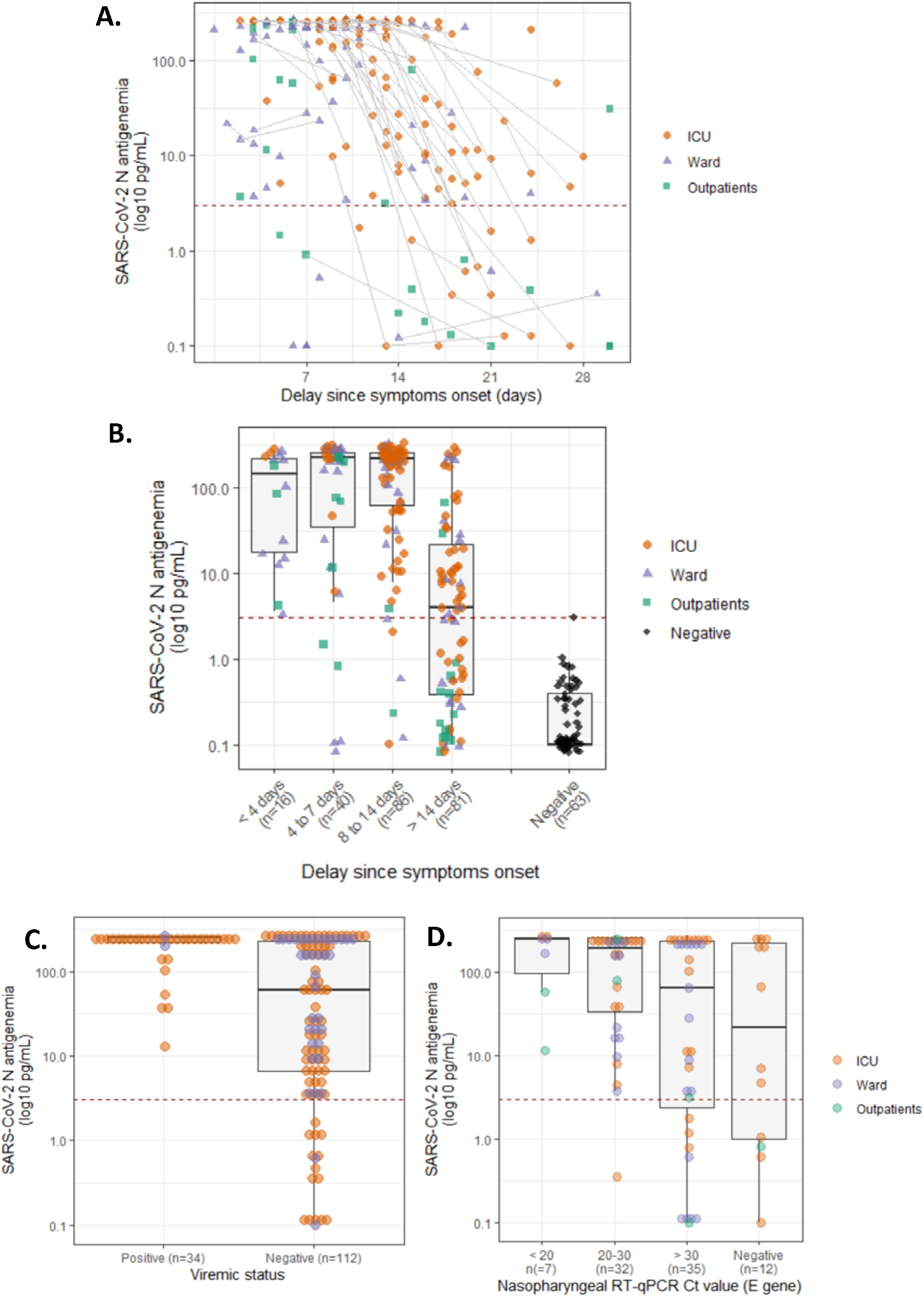
(A) Evolution of N-antigen sera levels in SARS-CoV-2-infected patients according to hospitalization status (N = 227 serum samples from 167 patients); sequential samples are connected with a gray line, while the positivity threshold value for N-antigen (2.97 pg/mL) is indicated with a dashed red line. (B) N-antigenemia levels according to delay since symptoms onset. (C) N-antigen sera levels according to positive and negative RNAaemia status (N = 85 patients). (D) N-antigen sera levels according to E-gene cycle threshold value of nasopharyngeal swabs collected within 24 hours (N = 73 patients).

In a subset of 73 COVID-19 patients with a serum collected within 24 hours following or preceding a positive nasopharyngeal swab using the RealStar® SARS-CoV-2 RT-PCR Kit (Altona, Germany) or the Cobas® SARS-CoV-2 Test (Roche, Switzerland), N-antigen detection was compared to the E gene cycle threshold value (Figure 1-D). Among patients with a Ct value below 30 and 33 on the nasopharyngeal swab, only 1/50 and 4/67 tested negative for N-antigenemia, respectively (Supplementary Appendix 1B). The former patient was sampled at day 21 following symptoms onset. Among patients with a negative nasopharyngeal RT-PCR, 8/12 presented positive N-antigenemia. The lower respiratory tract was explored for 6 out of these 8 patients at the same time point or within 5 days after, showing positive PCR in 5 cases.

## DISCUSSION

To our knowledge, this is the first demonstration of N-antigen blood accumulation during COVID-19. The assay evaluated in this work presented sensitivity above 90% during the acute phase of the disease *(i.e*. < 14 days after symptoms onset in PCR confirmed COVID-19 patients). N-antigen negativity was associated with anti-N IgG detection (6/10) and low nasopharyngeal viral load in the same 24 hours (7/7, > 30 Ct). These performances seems in line with PCR sensitivity reporting sensitivity rates of between 71 and 98%, based on negative RT-PCR tests which were positive on repeat testing^6^.

A limitation of this first N-antigenemia evaluation for COVID-19 diagnosis is the high proportions of ICU and ward patients. N-antigenemia decrease seems to occur earlier in outpatients, but have to be confirmed in larger cohorts.

This innovative marker opens new perspectives for diagnostic, such as rapid antigen blood test and combined ELISA assays, detecting both antigens and antibodies. This could provide a valuable new option for COVID-19 diagnosis, only requiring a blood draw, scalable in all clinical laboratories and high throughput analyzers.

## Data Availability

A file compiling all data used in this article is available on Mendeley Data public repository (https://data.mendeley.com/datasets/fjz6zbkxvm/1).

## ACKNOWLEDGEMENT

The French COVID cohort was sponsored by Inserm. We wish to thank the ANRS (Agence Nationale de la Recherche sur le SIDA et les hépatites virales).

## The members of the French COVID cohort management Committee study group (by alphabetical order)

Alpha DIALLO, Christelle PAUL, Noémie MERCIER, Soizic LE MESTRE, Ventzislava PETROV-SANCHEZ (ANRS, Paris, France), Claire ANDREJAK (CHU Amiens, France), Denis MALVY (CHU Bordeaux, France), Dominique DEPLANQUE, François DUBOS (CHU Lille, France), Patrick ROSSIGNOL (CHU Nancy, France), Manuel ETIENNE (CHU Rouen, France), Bénédicte ROSSIGNOL, Claire LEVY-MARCHAL, Tristan GIGANTE (FCRIN INI-CRCT, Paris, France), BELUZE Marine (F-CRIN Partners Platform, Paris, France), Anissa CHAIR, Antoine KHALIL, Benoit VISSEAUX, Camille COUFFIGNAL, Carine ROY, Cédric LAOUÉNAN, Charlene DA SILVEIRA, Coralie TARDIVON, Delphine BACHELET, Diane DESCAMPS, France MENTRÉ, François BOMPART, François-Xavier LESCURE, Gilles PEYTAVIN, Isabelle GORENNE, Isabelle HOFFMANN, Jade GHOSN, Jean Christophe LUCET, Jean-François TIMSIT, Jimmy MULLAERT, Krishna BHAVSAR, Lila BOUADMA, Lysa TAGHERSET, Marie-Capucine TELLIER, Marie-Pierre DEBRAY, Marina ESPOSITO-FARESE, Marion SCHNEIDER, Minh LE, Nadia ETTALHAOUI, Nassima SI MOHAMMED, Nathalie GAULT, Nathan PEIFFER-SMADJA, Ouifiya KEFIF, Philippine ELOY, Quentin LE HINGRAT, Samira LARIBI, Sarah TUBIANA, Théo TREOUX, Xavier DUVAL (Hôpital Bichat, Paris, France), Noémie VANEL (hôpital la timone, Marseille, France), Olivier PICONE, Romain BASMACI (Hôpital Louis Mourier, Colombes, France), François ANGOULVANT (Hôpital Necker, Paris, France), Florentia KAGUELIDOU, Justine PAGES (Hôpital Robert Debré, Paris, France), Aurélie VEISLINGER, Christelle TUAL (Inserm CIC-1414, Rennes, France), Aurélie PAPADOPOULOS, Hélène ESPEROU, Salma JAAFOURA, Sandrine COUFFIN-CARDIERGUES (Inserm sponsor, Paris, France), Mireille CARALP (Inserm Transfert, Paris, France), Alexandra COELHO, Alexandre HOCTIN, Alphonsine DIOUF, Marina MAMBERT (Inserm UMR 1018, Paris, France), Alexandre GAYMARD, Bruno LINA, Manuel ROSA-CALATRAVA, Maude BOUSCAMBERT, Olivier TERRIER (Inserm UMR 1111, Lyon, France), Amina MEZIANE, Céline DORIVAL, Dehbia BENKEROU, François TÉOULÉ (Inserm UMR 1136, Paris, France), Guillaume LINGAS, Hervé LE NAGARD, Jérémie GUEDJ, Nadège NEANT (Inserm UMR 1137, Paris, France), Laurent ABEL (Inserm UMR 1163, Paris, France), Coralie KHAN, Mathilde DESVALLÉE (Inserm UMR 1219, Bordeaux, France), Aurélie WIEDEMANN, Yves LEVY (Inserm UMR 955, Créteil, France), Hugo MOUQUET, Sylvie BEHILILL, Sylvie van der WERF, Vincent ENOUF (Pasteur Institute, Paris, France), Caroline SEMAILLE, Eric d’ORTENZIO, Minerva CERVANTES-GONZALEZ, Oriane PUÉCHAL (REACTing, Paris, France), Marion NORET (RENARCI, Annecy, France).

## The member of the CoV-CONTACT study group

### Principal investigator

DUVAL Xavier;

### Steering Committee

BURDET Charles, DUVAL Xavier, LINA Bruno, TUBIANA Sarah, VAN DER WERF Sylvie;

### CoV-CONTACT Clinical Centers

ABAD Fanny, ABRY Dominique, ALAVOINE Loubna, ALLAIN Jean-Sébastien, AMIEL-TAIEB Karline, AUDOIN Pierre, AUGUSTIN Shana, AYALA Sandrine, BANSARD Hélène, BERTHOLON Fréderique, BOISSEL Nolwenn, BOTELHO-NEVERS Elisabeth, BOUILLER Kévin, BOURGEON Marilou, BOUTROU Mathilde, BRICK Lysiane, BRUNEAU Léa, CAUMES Eric, CHABOUIS Agnès, CHAN THIEN Eric, CHIROUZE Catherine, COIGNARD Bruno, COSTA Yolande, COSTENOBLE Virginie, COUR Sylvie, CRACOWSKI Claire, CRACOWSKI Jean Luc, DEPLANQUE Dominique, DEQUAND Stéphane, DESILLE-DUGAST Mireille, DESMARETS Maxime, DETOC Maelle, DEWITTE Marie, DJOSSOU Felix, ECOBICHON Jean-Luc, ELREZZI Elise, FAUROUS William, FORTUNA Viviane, FOUCHARD Julie, GANTIER Emilie, GAUTIER Céline, GERARDIN Patrick, GERSET Sandrine, GILBERT Marie, GISSOT Valérie, GUILLEMIN Francis, HARTARD Cédric, HAZEVIS Béatrice, HOCQUET Didier, HODAJ Enkelejda, ILIC-HABENSUS Emila, JEUDY A, JEULIN Helene, KANE Maty, KASPRZYK Emmanuelle, KIKOINE John, LAINE Fabrice, LAVIOLLE Bruno, LEBEAUX David, LECLERCQ Anne, LEDRU Eric, LEFEVRE Benjamin, LEGOAS Carole, LEGRAND Amélie, LEGRAND Karine, LEHACAUT Jonathan, LEHUR Claire, LEMOUCHE Dalila, LEPILLER Quentin, LEPUIL Sévérine, LETIENNE Estelle, LUCARELLI Aude, LUCET Jean-Christophe, MADELINE Isabelle, MAILLOT Adrien, MALAPATE Catherine, MALVY Denis, MANDIC Milica, MARTY-QUINTERNET Solène, MEGHADECHA Mohamed, MERGEAY-FABRE Mayka, MESPOULHE Pauline, MEUNIER Alexandre, MIGAUD Maria-Claire, MOTIEJUNAITE Justina, GAY Nathalie, NGUYEN Duc, OUBBEA Soumaya, PAGADOY MaÏder, PARIS Adeline, PARIS Christophe, PAYET Christine, PEIFFER-SMADJA Nathan, PEREZ Lucas, PERREAU Pauline, PIERREZ Nathalie, PISTONE Thierry, POSTOLACHE Andreea, RASOAMANANA Patrick, REMINIAC Cécile, REXAH Jade, ROCHE-GOUANVIC Elise, ROUSSEAU Alexandra, SCHOEMAECKER Betty, SIMON Sandrine, SOLER Catherine, SOMERS Stéphanie, SOW Khaly, TARDY Bernard, TERZIAN Zaven, THY Michael, TOURNIER Anne, TYRODE Sandrine, VAUCHY Charline, VERDON Renaud, VERNET Pauline, VIGNALI Valérie, WAUCQUIER Nawal;

### Coordination and statistical analyses

BURDET Charles, DO THI THU Huong, LAOUÉNAN Cédric, MENTRE France, PAULINE Manchon, TUBIANA Sarah, DECHANET Aline, LETROU Sophie, QUINTIN Caroline, FREZOULS Wahiba;

### Virological lab

LE HINGRAT Quentin, HOUHOU Nadhira, DAMOND Florence, DESCAMPS Dianes, CHARPENTIER Charlotte, VISSEAUX Benoit, VABRET Astrid, LINA Bruno, BOUSCAMBERT Maud, VAN DER WERF Sylvie, BEHILLIL Sylvie, GAILLANNE Laurence, BENMALEK Nabil, ATTIA Mikael, BARBET Marion, DEMERET Caroline, ROSE Thierry, PETRES Stéphane, ESCRIOU Nicolas, BARBET Marion, PETRES Stéphane, ESCRIOU Nicolas, GOYARD Sophie;

### Biological center

KAFIF Ouifiya, PIQUARD Valentine, TUBIANA Sarah;

### Partners

RECOVER, REACTING, Santé Publique France (COIGNARD Bruno, MAILLES Alexandra), Agences régionales de santé (SIMONDON Anne, DREYERE Marion, MOREL Bruno, VESVAL Thiphaine);

### Sponsor

Inserm, AMAT Karine, AMMOUR Douae, AQOURRAS Khadija, COUFFIN-CADIERGUES Sandrine, DELMAS Christelle, DESAN Vristi, DOUTE Jean Michel, ESPEROU Hélène, HENDOU Samia, KOUAKAM Christelle, LE MEUT Guillaume, LEMESTRE Soizic, LETURQUE Nicolas, MARCOUL Emmanuelle, NGUEFANG Solange, ROUFAI Layidé;

### Genetic

LAURENT Abel, CAILLAT-ZUCMAN Sophie.

